# Outcomes of low-risk non-gynecological abdominal surgery in pregnant and nonpregnant women: a comparative study

**DOI:** 10.1101/2025.01.29.25321325

**Authors:** Abdulrahman Muaod Alotaibi

**Affiliations:** Associate Professor, Department of Surgery, Faculty of Medicine, University of Jeddah, Jeddah, Saudi Arabia; Department of Surgery, Dr. Soliman Fakeeh Hospital, Jeddah, Saudi Arabia

**Keywords:** acute appendicitis, pregnancy, laparoscopic appendectomy, laparoscopic cholecystectomy, biliary disease, cohort study

## Abstract

Women typically undergo an appendectomy and cholecystectomy, which are the most commonly performed non-gynecological abdominal surgeries. Despite advancements, research on the safety and efficacy of laparoscopic surgery for low-risk, non-gynecological abdominal conditions in later-stage pregnant women remains limited. This study aimed to assess the efficacy and postoperative outcomes of low-risk non-gynecological abdominal surgery in female patients, including pregnant women. Female patients diagnosed with appendicitis and biliary disease were included. Based on the pregnancy status, the participants were divided into pregnant and nonpregnant groups. The study identified 11 (1%) cases of pregnancy among the 1029 female patients enrolled. Patients in the pregnant group were younger (30 ± 4.7 years vs. 39 ± 13.5 years, p = 0.002), more likely to undergo emergency surgery (90 vs. 21%, p = 0.001), had a higher rate of undergoing the open-entry approach (18.2 vs. 2.1%, p = 0.002), and more frequently required a postoperative drain (36.4 vs. 10.4%, p = 0.023) than did those in the nonpregnant group. The comorbidities, American Society of Anesthesiologists score, length of hospital stay, operative time, blood loss, postoperative complications, 30-day readmission rate, and mortality rate remained statistically consistent across both groups. Surgery was performed primarily for acute appendicitis in 72% of pregnant women and for biliary disease in 83% of nonpregnant women. None of the pregnant patients experienced preterm labor or fetal mortality after surgery. One patient had an abortion 8 weeks after laparoscopic appendectomy. In conclusion, the overall complication rate of non-gynecological abdominal surgery for appendicitis and biliary disease is 2%. Biliary disease is the most common indication of non-gynecological abdominal surgery among nonpregnant women, whereas appendicitis is the most common indication among pregnant women. Emergency surgery is a significant predictor of complications in pregnant women.

## Introduction

The most common reasons for gynecological surgery in female patients are potentially torsioned ovaries and the presence of symptomatic masses in the adnexal area. Conversely, women undergo non-gynecological abdominal surgery for cholecystitis and acute appendicitis [1]. The laparoscopic approach can minimize postoperative pain, time for recovery, ileus and wound infections, inpatient expenditure, hospitalization, and thromboembolic events [2]. There is a minimal disparity in the success rates of laparoscopic abdominal surgery (LAS) between male and female patients. The safety and effectiveness of LAS in pregnant women have received special attention. Approximately 2% of pregnant women undergo abdominal surgeries, the most common of which are appendectomies and cholecystectomies [3]. Laparoscopy is generally safe but is associated with a higher rate of miscarriage than is laparotomy; therefore, choosing between the two is crucial [3].

Pregnant women face distinctive difficulties owing to physiological changes during pregnancy and concerns about how LAS could affect both the mother and baby. Consequently, pregnant patients have been prioritized in surgical care as well as in recent research on LAS. Laparoscopic cholecystectomy (LC) is a new obstetric procedure that was first performed in 1991. To reduce the risk of complications for the mother and fetus during an open-approach surgery, the introduction of minimally invasive surgery for pregnant women is a significant advancement in surgical care. Such advancements have helped adopt laparoscopic surgery for obstetric patients with multiple conditions [4]. Laparoscopy during pregnancy has been a polemic topic owing to concerns regarding fetal anomalies, miscarriages, and premature labor. However, a growing body of research over the last 20 years has shown that laparoscopic procedures have been safe and effective for pregnant women [5,6]. Consequently, laparoscopy during pregnancy has gained widespread acceptance. However, the research on laparoscopic surgery during pregnancy, particularly in the second and third trimesters, is still in its infancy. Surgeons have historically been hesitant to perform laparoscopic procedures on pregnant women owing to concerns regarding fetal safety and potential complications. This has resulted in a conservative approach, often delaying surgery until the second trimester, which is considered the optimal period for such interventions [7]. The available evidence is frequently of weak-to-moderate quality, with a limited number of prospective randomized trials undertaken in this field. Most available data were derived from retrospective studies, case reports, and systematic reviews, thereby constraining the robustness of the conclusions. Numerous studies have included small sample sizes, highlighting the deficiencies in large-scale multicenter trials. One study examined 23 patients over an 11-year period and highlighted the infrequency and under-reporting of these procedures [8]. A systematic review encompassing 590 patients from 51 studies highlighted the fragmented nature of the existing data [9]. The guidelines and practices for laparoscopic surgery during pregnancy have developed over time. Previous guidelines were more conservative; however, recent reviews and guidelines have questioned these established beliefs [7]. Therefore, this study aimed to validate the safety and feasibility of laparoscopic surgery during the second and third trimesters of pregnancy by contesting the conventional perspective that restricts surgery to the second trimester. This study highlights the benefits of a reduced duration of hospital stay and lower complication rates, thereby enhancing maternal and fetal outcomes. This study is believed to enhance the clinical guidelines, promote early diagnosis and intervention, and provide insights into the technical aspects of laparoscopic surgery in pregnant patients.

## Materials and methods

Patients were retrospectively evaluated, and data were collected from 1 January 2016 to 31 December 2019 at Dr. Soliman Fakeeh Hospital in Jeddah, Saudi Arabia. The inclusion criteria were designed to focus on common non-gynecological conditions requiring abdominal surgery and involving a low-risk profile, making them suitable for elective or emergency laparoscopic procedures.

The inclusion criteria of this study were as follows: female patients older than 18 years undergoing elective or emergency non-gynecological low-risk abdominal surgery, specifically appendectomy and cholecystectomy. The disease conditions being treated included appendicitis, biliary colic, obstructive jaundice, cholecystitis, and biliary pancreatitis. Patients undergoing gynecological procedures and those with trauma and diagnoses outside the scope of the inclusion criteria were excluded.

The patients were categorized into two groups: pregnant and nonpregnant. The diagnosis and decision for a surgical intervention were based on a combination of clinical evaluation, ultrasonography, magnetic resonance imaging, and diagnostic laparoscopy. The Institutional Review Board of the Fakeeh College for Medical Science approved the study protocol (approval number 200/IRB/2021 and 228/IRB/2021). The Institutional Review Board of the Fakeeh College for Medical Science waived the need for informed consent due to the retrospective nature of the study.

The primary outcome of this study was the safety and efficacy of laparoscopic surgery in both pregnant and nonpregnant women, defined by the absence of major intraoperative events such as fetal distress, premature labor, abortion, or 30-day postoperative complications (e.g., bleeding, conversion to open surgery, and infection). Secondary outcomes included the duration of hospital stay, defined as the time from the date of surgery to discharge; this was measured to determine whether pregnant patients had more extended stays than did nonpregnant patients. Adverse pregnancy outcomes were defined as the incidence of miscarriage, preterm labor, or adverse fetal outcomes. The research register in https://www.researchregistry.com with ID 10931.

### Surgical technique

Owing to their minimal invasiveness and good results, laparoscopic appendectomy (LA) and LC are the preferred methods for treating appendiceal and biliary diseases at our medical center. The surgeon’s preference and patient’s condition should be considered when determining whether to use a laparoscopic, open, or conversion approach. Factors such as the disease severity, patient’s comorbidities, surgical history, and intraoperative findings may influence the choice of the surgical technique to ensure the best possible outcome for each case.

### Statistical analysis

The analysis included 16 variables. Continuous variables are presented as means and standard deviations. The median and interquartile ranges were calculated for non-normally distributed continuous variables. The normality of the data was evaluated using the Shapiro– Wilk test. Continuous variables exhibiting a normal distribution were analyzed using the unpaired Student’s t-test.

By contrast, the Mann–Whitney U test was used to analyze the variables that did not follow a normal distribution. Fisher’s exact test was used to compare the surgical outcomes between pregnant and nonpregnant women. A multivariate logistic regression model was constructed twice, first by including all variables and then by using the variables found to be significant in the bivariate analysis to identify the predictors of postoperative complications, adjusting for potential confounders. A two-sided p-value of less than 0.05 was considered statistically significant. All data analyses were conducted using SPSS software (version 25, SPSS Inc., Chicago, IL, USA).

## Results

### Age distribution of the study participants

The mean (standard deviation [SD]) age of the study participants was 38.97 (13.52) years. When stratified by pregnancy status, the mean (SD) age of nonpregnant women was 39.06 (13.56) years, whereas that of pregnant women was 30.36 (4.72) years. The age distribution between the two groups was statistically significant (p = 0.015), indicating a significant difference in the ages of pregnant and nonpregnant participants. The age distribution of the participants is shown in Fig 1.

**Fig 1.**
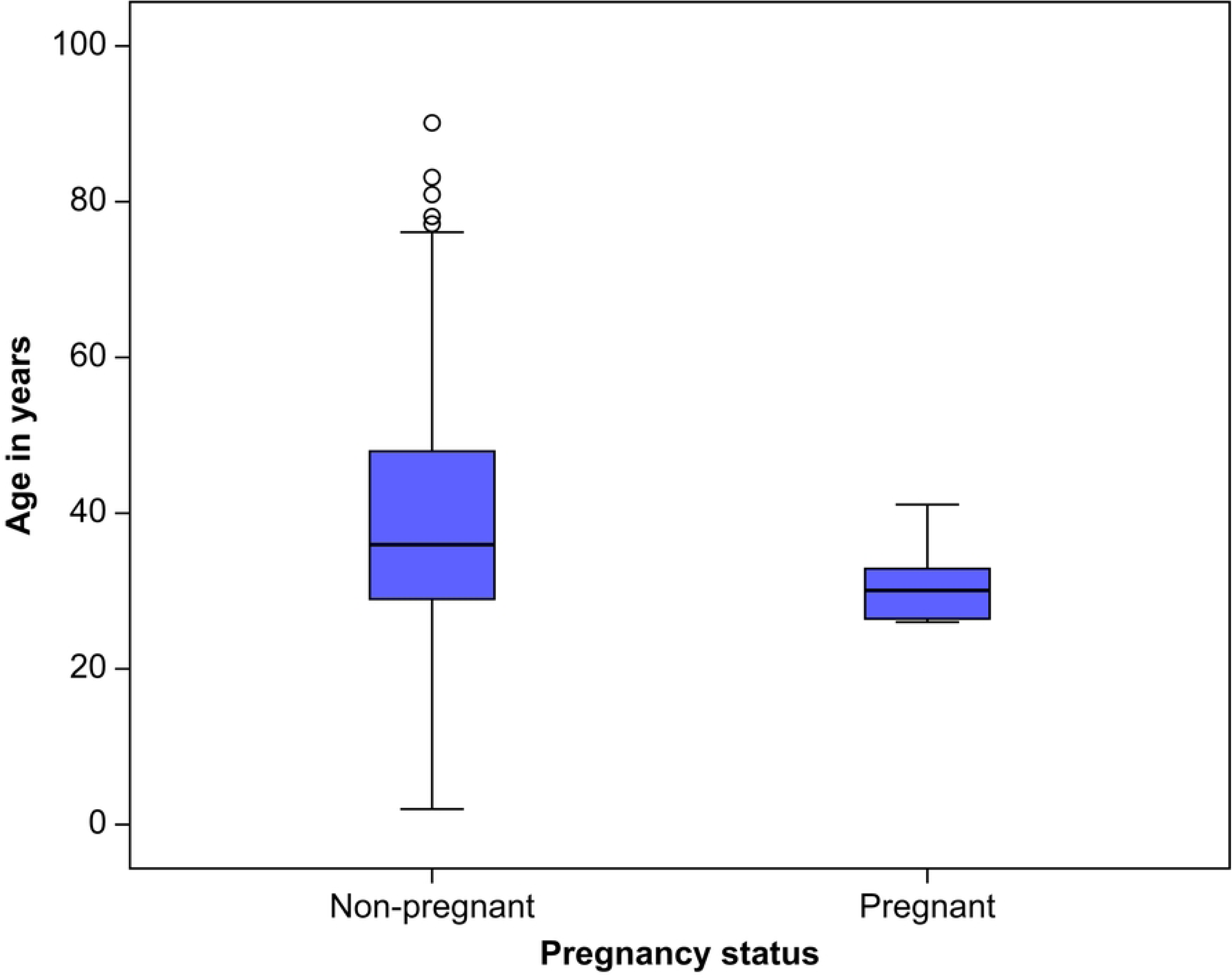
Age distribution of the study participants.

### Characteristics of pregnant women

A total of 11 pregnant women were included in the study. The mean (SD) gestational age was 15.55 (7.79) weeks, with a median (interquartile range) of 13 (8–24) weeks. Among the pregnant women, six (54.5%) were in their first trimester, four (36.4%) were in their second trimester, and one (9.1%) was in their third trimester. The distribution of pregnant women according to their trimester and diagnoses is presented in Table 1.

**Table 1.** Distribution of Pregnant Women According to Their Trimester (n = 11) and Diagnosis.

| **Pregnancy in Weeks** | **Cholecystitis (n=2)** | **Pancreatitis (n=1)** | **Acute Appendicitis (n = 8)** | **p-value<sup>a</sup>** |
| --- | --- | --- | --- | --- |
|  |  |  |  | *0.35* |
| **First trimester** |  |  |  |  |
| **6** |  |  | 1 |  |
| **8** |  | 1* | 1 |  |
| **10** |  |  | 1 |  |
| **11** | 1 |  |  |  |
| **13** |  |  | 1 |  |
| **Second trimester** |  |  |  |  |
| **17** |  |  | 1 |  |
| **21** | 1 |  |  |  |
| **24** |  |  | 1 |  |
| **26** |  |  | 1 |  |
| **Third trimester** |  |  |  |  |
| **27** |  |  | 1 |  |

<sup>a</sup>Fisher's exact test
\*Missed abortion

### Disease status

Cholecystitis was the most common diagnosis among nonpregnant women and was found in 506 (49.7%) patients. By contrast, uncomplicated appendicitis was the most prevalent condition among pregnant women and was noted in six (54.5%) patients. Fig 2 and Table 2 presents the diagnoses of patients in the two groups of this study.

**Fig 2.**
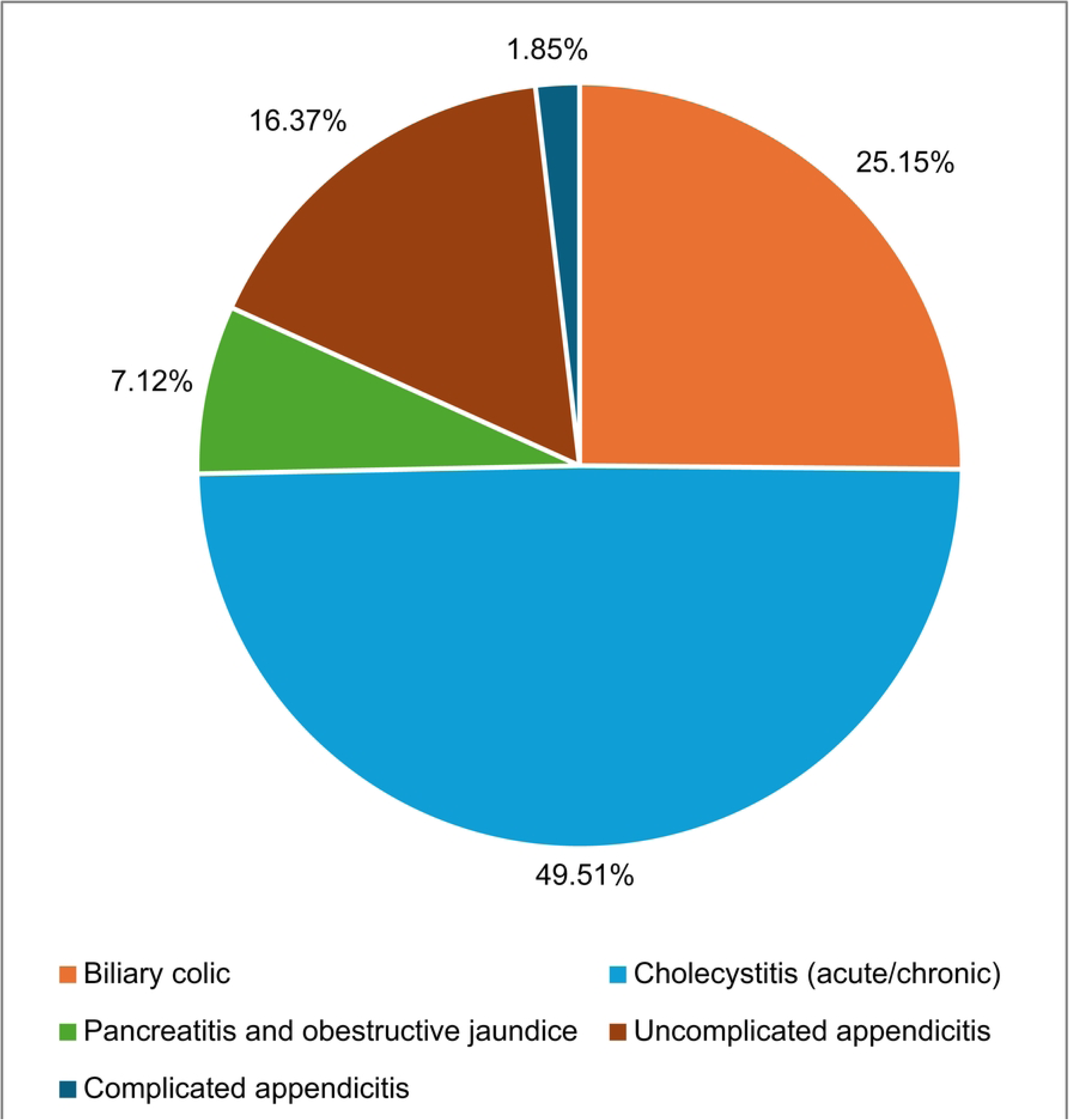
Clinical diagnosis of 1029 female patients.

**Table 2.** Disease Conditions of the Study Participants.

| S.<br>No | Disease Condition | Nonpregnant<br>Women | Pregnant<br>Women | p-value |
| --- | --- | --- | --- | --- |
|  |  | Number (%) | Number (%) |  |
| 1 | Cholecystitis | 508 (99.6) | 2 (0.4) |  |
| 2 | Biliary colic | 259 (100) | 0 (0) |  |
| 3 | Non-complicated appendicitis | 162 (96.4%) | 6 (3.6%) |  |
| 4 | Pancreatitis and obstructive<br>jaundice | 72 (98.6%) | 1 (1.4%) | 0.001* |
| 5 | Complicated appendicitis | 17 (89.5%) | 2 (10.5%) |  |

\*p-value is significant

### Comparison of surgical outcomes between nonpregnant and pregnant women

Among the 1029 female patients enrolled in the study, 11 (1%) were identified as pregnant. Participants in the pregnant group were younger (mean age: 30 ± 4.7 years vs. 39 ± 13.5 years, p = 0.002), more likely to undergo emergency surgery (90% vs. 21%, p = 0.001), had a higher rate of undergoing the open surgical approach (18.2% vs. 2.1%, p = 0.002), and more frequently required a postoperative drain (36.4% vs. 10.4%, p = 0.023) than did those in the nonpregnant group. However, the two groups showed no statistically significant differences in terms of comorbidities, the American Society of Anesthesiologists score, length of hospital stay, operative time, blood loss, postoperative complications, 30-day readmission, or mortality rates (Table 3).

**Table 3.** Demographics and Clinical Features of the Study Participants According to Their Pregnancy Status.

| **Variables** | **Nonpregnant Group** | **Pregnant Group** | **p-value<sup>a</sup>** |
| --- | --- | --- | --- |
|  | **(n = 1018)** | **(n = 11)** |  |
| **Age at the time of surgery, years** | 39 ± 13.5 | 30 ± 4.7 | **0.002\*** |
| **ASA score** |  |  | 0.88 |
| I–III | 1016 (99.8) | 11 (100) |  |
| IV–V | 2 (0.2) | 0 (0) |  |
| **Length of hospital stay** |  |  | 0.36 |
| ≤72 h | 923 (90.7) | 9 (81.8) |  |
| >72 h | 95 (9.3) | 2 (18.2) |  |
| **Comorbidities** |  |  | 0.27 |
| Absent | 576 (56.6) | 8 (72.7) |  |
| Present | 442 (43.4) | 3 (27.3) |  |
| **Clinical diagnosis** |  |  |  |
| Biliary colic | 259 (25.4) | 0 (0) |  |
| Cholecystitis (acute/chronic) | 508 (49.9) | 2 (18.2) |  |
| Pancreatitis/obstructive jaundice | 72 (7.1) | 1 (9.1) |  |
| Non-complicated appendicitis | 162 (16.0) | 6 (54.5) |  |
| Complicated appendicitis | 17 (1.6) | 2 (18.2) |  |
| **Surgical setting** |  |  | **0.001\*** |
| Elective | 803 (78.9) | 1 (9.1) |  |
| Urgent/ER | 215 (21.1) | 10 (90.9) |  |
| **Surgical approach** |  |  | **0.002\*** |
| Laparoscopy | 989 (97.1) | 9 (81.8) |  |
| open | 21 (2.1) | 2 (18.2) |  |
| Laparoscopy converted to open | 8 (0.8) | 0 (0) |  |
| **Operative time (min)** | 53 ± 27.5 | 61.8 ± 28.1 | *0.29* |
| **Blood loss** |  |  | *0.93* |
| Yes (>100 mL) | 6 (0.6) | 0 (0) |  |
| No | 1012 (99.4) | 11 (100) |  |
| **Drain** |  |  | ***0.023\**** |
| Yes | 106 (10.4) | 4 (36.4) |  |
| No | 912 (89.6) | 7 (63.6) |  |
| **Postoperative complications** |  |  | *0.87* |
| Yes | 20 (2.0) | 2 (18.1) |  |
| No | 998 (98.0) | 9 (81.9) |  |
| **30-day readmission** |  |  | *0.17* |
| Yes | 16 (1.6) | 1 (9.1) |  |
| No | 1002 (98.4) | 10 (90.9) |  |
| **Abortion** | - | 1 |  |
| **30-day mortality** | 0 | 0 |  |

Values are expressed as means ± standard deviation or as number of patients (%).
<sup>a</sup>Fisher's exact test
\*p-value is significant
SD, standard deviation; ASA, American Society of Anesthesiologists; ER, emergency

The mean (SD) operative time for nonpregnant women was 52.95 (27.47) min, whereas that for pregnant women was 61.82 (28.06) min. Although the Mann–Whitney U test indicated that pregnant women required longer surgery time than did nonpregnant women, this difference was not statistically significant (p = 0.184). One missed abortion in this group occurred in the first trimester, and one case of wound infection occurred later in pregnancy.

### Multivariate logistic regression model

The variables of surgical setting, surgical procedures, and postoperative drainage were found to be significant in the bivariate analysis. These variables were subsequently included in the multivariate logistic regression analysis to develop a predictive model. The results of the multivariate logistic regression analysis are presented in Table 4. After adjusting for confounding factors, the results indicated that only one variable—the surgical setting—emerged as a significant predictor of surgical complications in pregnant women, with an odds ratio (OR) of 32.13 (95% confidence interval, 4.02–256.79; p = 0.001). Therefore, the setting in which the surgery is performed (e.g., emergency vs. elective) notably affects the likelihood of surgical complications during pregnancy.

**Table 4.** Multivariate Logistic Regression Model for the Surgical Outcomes of Study Participants.

| S. No. | Surgical Outcomes | Adjusted OR (95% CI) | p-value |
| --- | --- | --- | --- |
| 1 | Operation setting | 32.13 (4.02–256.79) | 0.001* |
| 2 | Surgical approach | 1.225 (0.34–4.39) | 0.756 |
| 3 | Drain | 3.02 (0.83–10.92) | 0.093 |

The value of the constant is -10.527
\*p-value is significant
OR, odds ratio; CI, confidence interval

## Discussion

In this cohort, the total complication rate for non-gynecological abdominal surgeries, such as those conducted for appendicitis and biliary disorders, is roughly 2%. the leading cause of abdominal surgery in nonpregnant and pregnant women is biliary disease and appendicitis, respectively. The need for surgical intervention has a significant impact on results; the risk of complications for pregnant patients is substantially higher with emergency surgeries compared to elective procedures. The conventional treatments for appendicitis and symptomatic cholelithiasis are LA and LC, respectively. Minimally invasive surgeries are now widely accepted as the preferred care method because of their advantages such as decreased postoperative discomfort, quicker recovery periods, reduced infection risks, and shorter hospital stays than those associated with traditional open surgical methods [10]. Appendicitis is suspected in 1 of 600 pregnancies annually and is the leading indication for non-obstetric surgery during pregnancy [11,12]. A higher percentage of women experience appendicitis during the second trimester of pregnancy than during the first and third trimesters. The increased susceptibility to appendicitis during the second trimester of pregnancy can be explained by several factors. The appendix moves out of its normal position because of the pressure exerted by the expanding uterus. Owing to this displacement, the appendix may become more sensitive to irritation and inflammation. In addition, the appendiceal blood supply could be jeopardized because the expanding uterus increases the intra-abdominal pressure. Altered blood flow, combined with increased intraluminal pressure, can lead to appendiceal obstruction, which is a significant cause of appendicitis. In addition, the maternal immune system undergoes changes to tolerate the fetal growth. These modifications may slightly weaken the body’s ability to fight infections, increasing the risk of infections such as appendicitis. Together, these factors contribute to a higher likelihood of appendicitis during the second trimester of pregnancy than during the first and third trimesters [10,12,13]. When appendicitis is complicated, such as in cases of perforation or peritonitis, the rate of fetal loss significantly increases from 1.5% to an estimated rate of 30%. The reasons for increased fetal loss in complicated appendicitis include maternal sepsis, peritonitis, compromised fetal oxygen and nutrient supply, surgical stress associated with anesthesia and operative time, and delayed diagnosis and treatment because of atypical presentations [14,15]. In the first 3 months of pregnancy, some studies reported a fetal loss in 3– 20% of women who have had a problematic appendicectomy [16–18], with a 15–45% preterm delivery rate [19], as well as a higher risk of spontaneous abortion and fetal demise [20].

A study involving 3158 individuals from 632 acute care hospitals compared the outcomes of appendectomy with those of conservative management [21]. During the second trimester, appendectomy was associated with an increased incidence of preterm delivery or abortion (OR, 3.0). Antepartum hemorrhage was more prevalent after appendectomy in the first (OR, 2.0) and third (OR, 2.5) trimesters. The clinical outcomes of acute appendicitis during gestation differ according to the trimester. Considering the risks associated with appendectomy, conservative management may be appropriate, based on the clinical context and trimester [21]. The complication rate after appendectomy generally varies depending on factors such as the type of surgery (laparoscopic vs. open), severity of appendicitis (e.g., uncomplicated vs. perforated), and patient characteristics. The complication rate is typically low, ranging between 2% and 5% in uncomplicated cases [15]. This approach is associated with fewer wound infections, quicker recovery, and less postoperative pain than is open surgery. Complication rates can be high, particularly in cases of perforated appendicitis, with rates ranging from 5–10%. In severe cases (e.g., perforation or abscess), wound infection, intra-abdominal abscess, or bowel obstruction can occur [15].

In concordance with patient outcomes in this study, the use of antibiotics, close perioperative monitoring, interdisciplinary teamwork, and advancements in perioperative care have contributed to the significant reduction in both fetal and maternal mortality rates in cases of appendicitis and other non-obstetric surgeries during pregnancy [6,22–24].

Complex conditions such as perforated appendicitis, maternal sepsis, and fetal complications are best treated with antibiotics. Perioperative monitoring is necessary to quickly intervene in the event of distress. Optimal care is achieved through interdisciplinary teamwork, particularly for high-risk surgeries.

The results from a meta-analysis of 4694 patients disproved the hypothesis that LA increases the risk of fetal loss in pregnant women [24]. Conversely, other reports have indicated a potential increase in fetal complications using this approach. Certain studies have raised concerns that carbon dioxide insufflation used during laparoscopic procedures could negatively affect fetal perfusion or contribute to fetal acidosis, especially if the intra-abdominal pressure is not carefully monitored [25]. For example, in some analyses, the pooled risk of fetal loss following LA was noted to be higher than that following open appendectomy, with one meta-analysis suggesting a relative increase in fetal loss following LA, although the absolute risk remains low [24]. It is essential to consider that these risks may also be influenced by other factors, such as the timing of surgery (earlier vs. later in pregnancy), severity of appendicitis (uncomplicated vs. complicated), and skill and experience of the surgical team. Therefore, although the consensus is that laparoscopic surgery is safe during pregnancy, individual patient factors must always be considered to minimize the risk.

For LC, the complication rate is generally low at approximately 2–5%, making it the preferred method for gallbladder removal owing to faster recovery and fewer complications than those following open surgery [26].

The most common complications include bile duct injury (0.2–0.5%), bile leaks (0.3– 0.5%), wound infections (1–2%), bleeding, and abscess formation, which are rare. For open surgery, especially in complex cases, the complication rate is higher at approximately 5–10% [26].

Pregnant women exhibiting symptoms of cholelithiasis are typically counseled against surgical intervention because of the potential risks. Medical experts advocate prioritizing dietary modifications, pharmacological interventions, and vigilant monitoring until the postpartum period. This prudent strategy seeks to reduce gallbladder stimulation and the symptoms associated with gallstones, thereby securing optimal outcomes for both the mother and the infant [27].

Nevertheless, nonoperative management is associated with a substantial recurrence of symptoms, leading to heightened hospitalization rates and a 13% increase in pancreatitis associated with gallstones, which accounts for fetal loss in 10–20% of pregnancies [28–30]. For patients with acute cholecystitis (inflammation of the gallbladder), recurrent episodes of biliary colic, or complications such as gallstone pancreatitis, surgery may be necessary during pregnancy to prevent maternal and fetal harm. When surgery is required, LC is considered the standard of care and can be safely performed during all trimesters of pregnancy.

One of the patients in this study experienced biliary pancreatitis that was managed with stent placement followed by LC. Unfortunately, during treatment, the patient experienced a missed abortion at 8 weeks of gestation. This case highlights the potential risks associated with biliary tract diseases and their treatment during pregnancy, particularly when complications such as pancreatitis arise. Biliary pancreatitis during pregnancy is relatively rare, with an incidence rate ranging from 1 in 1000 to 1 in 4000 pregnancies [31]. The most common causes are gallstones and biliary sludge, which can obstruct the bile duct and cause pancreatic inflammation. Pregnant women, especially those in their second or third trimester, are at a higher risk owing to hormonal changes that slow down gallbladder emptying, thereby increasing the risk of gallstone formation.

A retrospective study in Beijing reported that, among 32 pregnant women with acute pancreatitis, 90% of the cases occurred during the third trimester, reflecting the increased risk with advancing gestational age. The primary causes are hypertriglyceridemia and gallstones, which are more common in Western populations [32].

Pregnant women who seek nonoperative treatment for gallbladder diseases, such as cholelithiasis or biliary pancreatitis, are at a risk of experiencing complications such as preterm birth, premature labor, or spontaneous abortions. Poor pregnancy outcomes can occur because of infection, inflammation, systemic effects caused by gallstones, or recurring biliary symptoms. By contrast, surgical removal of the gallbladder (cholecystectomy), mainly via laparoscopic techniques, reduces the risk of these adverse pregnancy outcomes when performed under controlled conditions. Studies have indicated that early surgical intervention to treat symptomatic gallstone disease, especially in cases of recurrent symptoms or complications such as biliary pancreatitis, can help mitigate the risks of preterm labor and spontaneous abortion. Nonoperative management tends to carry a higher risk of recurrent biliary symptoms, which can trigger preterm labor [32].

Immediate surgical intervention, when necessary, may reduce undesirable consequences in symptomatic patients. There is no need to wait until the second trimester to perform a laparoscopy, as some studies have shown that the procedure can be performed safely and effectively during the first trimester of pregnancy [7,33–35].

An examination of 18,630 pregnant patients who underwent cholecystectomy between 2016 and 2020 on a national scale showed that the rates of cholecystectomy in the second trimester versus the first and third trimesters were 55% versus 25% and 20%, respectively, and all groups had similar rates of pregnancy outcomes [36].

In one review, 10,632 patients from 11 comparative studies were included to assess the outcomes of laparoscopic versus open cholecystectomy in pregnant women. The average gestational age at which laparoscopic procedures were performed was 18 weeks, whereas open procedures were performed at 24 weeks. This was the first meta-analysis to show that LC was associated with significantly fewer maternal and fetal complications than was open cholecystectomy. The benefits of laparoscopic surgery include a lower incidence of complications such as preterm labor, spontaneous abortion, and fetal distress, primarily owing to its minimally invasive nature, reduced postoperative pain, quicker recovery, and lower infection rates [37]. This evidence supports the increasing preference for LC over open cholecystectomy in managing symptomatic gallbladder disease during pregnancy, given its safer profile for both the mother and fetus.

Independent predictors for complications and increased length of hospital stay after LC include a gallbladder wall thickness greater than 5 mm, an urgent and time-consuming operation, and postoperative drains [38].

Some researchers have linked uterine irritability to the onset of preterm labor after pregnancy-related surgery, but the exact cause of preterm birth after surgery remains unknown [32]. The present study found that the surgical setting is a significant predictor of surgical complications in pregnant women, as indicated by the results of multivariate regression analysis, even after adjusting for confounding variables. More specifically, preterm delivery is one of the complications associated with increased rates of emergency surgery, which induces an inflammatory response and makes the uterine muscles more sensitive, causing labor to start earlier than expected. Anxiety during emergency procedures, along with the immune and inflammatory reactions caused by surgical trauma, can worsen the mother’s health and increase the risk of complications during pregnancy. Multiple studies have found that planned surgical interventions that occur at the right time, rather than emergency procedures, improve outcomes for both the mother and unborn child [15].

The best outcome is ensured by accurate diagnosis using ultrasonography and magnetic resonance imaging [39] and by following the Society of American Gastrointestinal and Endoscopic Surgeons (SAGES) laparoscopic guidelines for pregnant women, which emphasize performing the procedure in the second trimester using an open-entry approach and maintaining the pressure of the pneumoperitoneum at 8–12 mmHg [31,40,41]. There were no reported access-related incidents between the two groups of women in the present study, even though 45% (5/11) of the operations were performed in the second and third trimesters, and the primary entry method was the Hasson technique.

In this study, the surgeon’s experience at a private tertiary center is presented, with the caution that a statistically significant age gap was observed between pregnant and nonpregnant women. Women in the pregnant group were younger than those in the nonpregnant group. In addition, the sample size of the pregnant group was relatively small, which should be considered when interpreting the results. Although these findings are consistent with the general tendencies observed in other studies of a similar nature, the small sample size of the pregnant cohort may limit the statistical power and generalizability of the results. Therefore, additional research should be conducted with larger sample sizes to confirm the effect of emergency setting on pregnant women and to concentrate on the possibility of converting an emergency surgery into an elective setting.

## Conclusions

Low-risk non-gynecological abdominal surgery is associated with a 2% postoperative complication rate in females, with the benefit of a shorter hospital stay. Acute appendicitis is a typical emergency that requires surgical intervention among pregnant women, whereas cholecystitis remains the most frequent diagnosis among nonpregnant women. Laparoscopy is safe throughout the pregnancy, although extra caution is necessary in cases of biliary pancreatitis during the first trimester when there is a risk of complications such as a missed abortion. Emergency surgery significantly predicts the complications that may arise during surgery, particularly in pregnant patients. In an emergency, the team must control the inflammatory response and reduce the stress caused by the surgical procedures to ensure favorable results. Possible research questions at various points in pregnancy include minimally invasive methods and methods for administering anesthesia that reduce the risk to the developing baby while alleviating the mother’s pain and inflammation.

It would be possible to determine the intervention thresholds by associating pre- and postoperative inflammatory marker monitoring with maternal and fetal outcomes in longitudinal studies. Preoperative counseling on the effects of stress on the mother and her recovery could improve psychosocial support for pregnant women undergoing surgery. Prospective studies that evaluate outcomes based on trimester-specific interventions could refine the guidelines for the safest timing of surgeries, including the role of conservative management and individualization of care.

## Data Availability

All relevant data are within the manuscript.

## Acknowledgments

Author would like to thank Editage.com for editing the manuscript.

